# Neural correlates of sleep recovery following melatonin treatment for pediatric concussion: a randomized control trial

**DOI:** 10.1101/2020.08.02.20166918

**Authors:** Kartik K. Iyer, Andrew Zalesky, Luca Cocchi, Karen M. Barlow

**Affiliations:** Child Health Research Centre, Faculty of Medicine, The University of Queensland, Brisbane, Australia; Melbourne Neuropsychiatry Centre & Department of Biomedical Engineering, The University of Melbourne, Australia; QIMR Berghofer Medical Research Institute, Clinical Brain Networks group, Australia; Department of Neurology, Queensland Children’s Hospital, Brisbane, Australia; Alberta Children’s Hospital Research Institute, Calgary, Canada; University of Calgary, Calgary, Canada

**Author notes:** Corresponding author: Dr Kartik K. Iyer, Corresponding author’s address: Child Health Research Centre, Faculty of Medicine, The University of Queensland, Brisbane, Australia., Corresponding author’s, Corresponding author’s.

**Keywords:** mild traumatic brain injury, concussion, brain connectivity, sleep, melatonin

## Abstract

Evidence-based treatments for children with persistent post-concussion symptoms (PPCS) are few and limited. Common PPCS complaints such as sleep disturbance and fatigue could be ameliorated via the supplementation of melatonin, which has significant neuroprotective and anti-inflammatory properties. This study aims to identify neural correlates of melatonin treatment with changes in sleep disturbances and clinical recovery in a pediatric cohort with PPCS. We examined structural and functional neuroimaging (MRI) in 62 children with PPCS in a randomized, double-blind, placebo-controlled trial of 3mg or 10mg of melatonin (NCT01874847). The primary outcome was the total youth self-report Post-Concussion Symptom Inventory (PCSI) score after 28 days of treatment. Secondary outcomes included the change in the sleep domain PCSI score and sleep-wake behavior (assessed using wrist-worn actigraphy). Whole-brain analyses of (i) functional connectivity (FC) of resting-state fMRI, and (ii) structural grey matter (GM) volumes via voxel-based morphometry were assessed immediately before and after melatonin treatment and compared to placebo in order to identify neural effects of melatonin treatment. Increased FC of posterior default mode network (DMN) regions with visual, somatosensory and dorsal networks was detected in the melatonin groups over time. FC increases also corresponded with reduced wake periods (*r=*−0.27, *p=*0.01). Children who did not recover (*n*=39) demonstrated significant FC increases within anterior DMN and limbic regions compared to those that did recover (i.e. PCSI scores returned to pre-injury level *n*=23) over time, (*p=*0.026). Increases in GM volume within the posterior cingulate cortex were found to correlate with reduced wakefulness after sleep onset (*r=*−0.32, *p=*0.001) and sleep symptom improvement (*r=*0.29, *p=*0.02). Although the melatonin treatment trial was negative and did not result in PPCS recovery (with or without sleep problems), the relationship between melatonin and improvement in sleep parameters were linked to changes in function-structure within and between brain regions interacting with the DMN.

## INTRODUCTION

Traumatic brain injuries (TBI) in children are one of the most common causes of neurological morbidity and mortality worldwide.^1^ Most children sustain a mild TBI (mTBI), and more than 25% of these children will develop persistent post-concussion symptoms (PPCS) at one-month post-injury.^2, 3^ Children with PPCS commonly experience a cluster of physical, psychological and behavioral symptoms. One of the most common complaints is the presence of sleep disturbances and difficulty maintaining regular sleep-wake rhythms.^4, 5^ The neural effects of these ongoing sleep disturbances in TBIs, which include insomnia, pleiosomnia, and fatigue, alongside headaches, dizziness and irritability common to PPCS are associated with changes in brain structure and function during recovery.^6-12^

In examining neural changes during pediatric PPCS recovery, fMRI connectivity studies have highlighted the significance of altered patterns of communications within brain regions comprising the default mode (DMN) and executive networks.^11-15^ In typically developing cohorts, sleep-related problems in children and adolescents are known to compromise the efficiency and connectivity of key nodes of the DMN, such as the posterior cingulate cortex (PCC).^16, 17^ Reports on changes to grey matter following PPCS, however, remain scarce. A recent pediatric study reported increased anisotropic diffusion in cortical and subcortical grey matter structures in thalamic and temporal cortices at 4-months post-injury.^8^ Though these findings highlight that changes in brain activity occur during recovery, little is known about the relationship of specific symptoms with structure-function alterations during recovery from PPCS. Adding to this literature, we recently reported that, in children with mTBI and PPCS, persisting sleep disturbance and fatigue are linked to decreases in functional connectivity and grey matter volumes within the posterior and anterior regions of the DMN one month following the injury.^9^ Given these evidences, the relationship between symptom resolution and brain network reorganization is likely to reveal important insights into the role of neuroplasticity and likelihood for recovery.

Pharmacological and non-pharmacological treatments that seek to the alleviate PPCS are limited, often yielding mixed results.^18^ The supplementation of neuroprotective agents such as melatonin could help ameliorate common sleep and fatigue complaints present in individuals with PPCS, and in turn, help reduce overall symptom burden and improve brain functions during recovery. Previous studies which have examined increased sleep disturbances in TBI cohorts^19-22^ have indicated that decreased melatonin production is strongly associated with reduced sleep efficiency, overall sleep quality and increased wake periods after sleep onset (WASO). Further studies in children and adolescents with sleep-wake disorders and related fatigue, have shown that melatonin treatment can improve sleep onset, increase sleep duration and reduce daytime sleepiness,^23^ whilst improving other physical complaints such as post-traumatic headaches following a head injury.^24^ However, evidences linking melatonin treatment with improved sleep and brain network response is less understood in TBI-related cohorts.

In consideration of existing evidences and prior work, the present study investigates the neural effects of melatonin as a treatment for children with PPCS and whether changes to anatomical and functional brain markers observed before and after treatment were associated with improvement in sleep parameters. Analyses of structural and functional MRI were performed in a randomized, double-blind, placebo-controlled clinical trial of melatonin in a pediatric PPCS cohort over a 4-week period, with neuroimaging obtained before and after treatment.^25^ A whole-brain connectomics approach was employed to ascertain treatment effects in conjunction with change in sleep parameters. This study hypothesized that, following melatonin treatment, associated changes in whole-brain grey matter and functional connectivity would enhance compensatory functions of the DMN and be positively associated with improvement in sleep parameters and overall recovery.

## METHODS

### Study design and participants

This study was a single-center, randomized, double-blind, placebo-controlled trial of melatonin administered in children (ages 8 to 18 years) with PPCS at 4 to 6 weeks post-injury, conducted at Alberta Children’s Hospital (NCT01874847) and has been reported previously.^25^ The clinical trial was conducted in accordance with Good Clinical Practice guidelines and enrollment occurred between February 2014 and April 2017. Participants were excluded if (1) they had had a previous concussion within 3-months; (2) a Glasgow Coma Scale (GCS) scores less than 13; (3) significant medical history including any subjects taking medications that were likely to affect neuroimaging and/or sleep; and (4) inability to complete questionnaires and/or neuropsychological evaluation. Participants who enrolled in the trial were also invited to participate in neuroimaging (MRI) scanning before and after treatment concurrent with completion of outcome measures. To ensure against sources of bias, participants who had neuroimaging did not significantly differ in age, sex, average household income, days post-injury, loss of consciousness at injury, pre or post-injury post-concussion symptom scores, or treatment response from trial participants who did not have neuroimaging performed. On enrollment, participants were randomized (1:1:1) into three treatment groups: (i) sublingual placebo (ii) sublingual melatonin 3 mg, and (iii) sublingual melatonin 10 mg. Participants, parents and investigators were blinded to treatment group. Melatonin dosage was administered daily for a 28-day period one hour before usual bedtime. The final cohort for this neuroimaging study included 62 children with PPCS (20 on placebo, 22 on 3mg melatonin, and 20 on 10 mg melatonin) with structural and resting-state functional MRI before and after treatment. Participants were not taking melatonin at the time of neuroimaging (i.e. 48-hour wash-out period; half-life of melatonin is ∼2 hours). **Figure 1** encapsulates study enrollment, randomization, neuroimaging and follow-up outcomes measured throughout the trial. Approval for this study was granted by the University of Calgary Conjoint Health Research Ethics Board (REB13-0372) and the University of Queensland (2017001523). Written and verbal parental consent and child assent was obtained.

**Figure 1.**
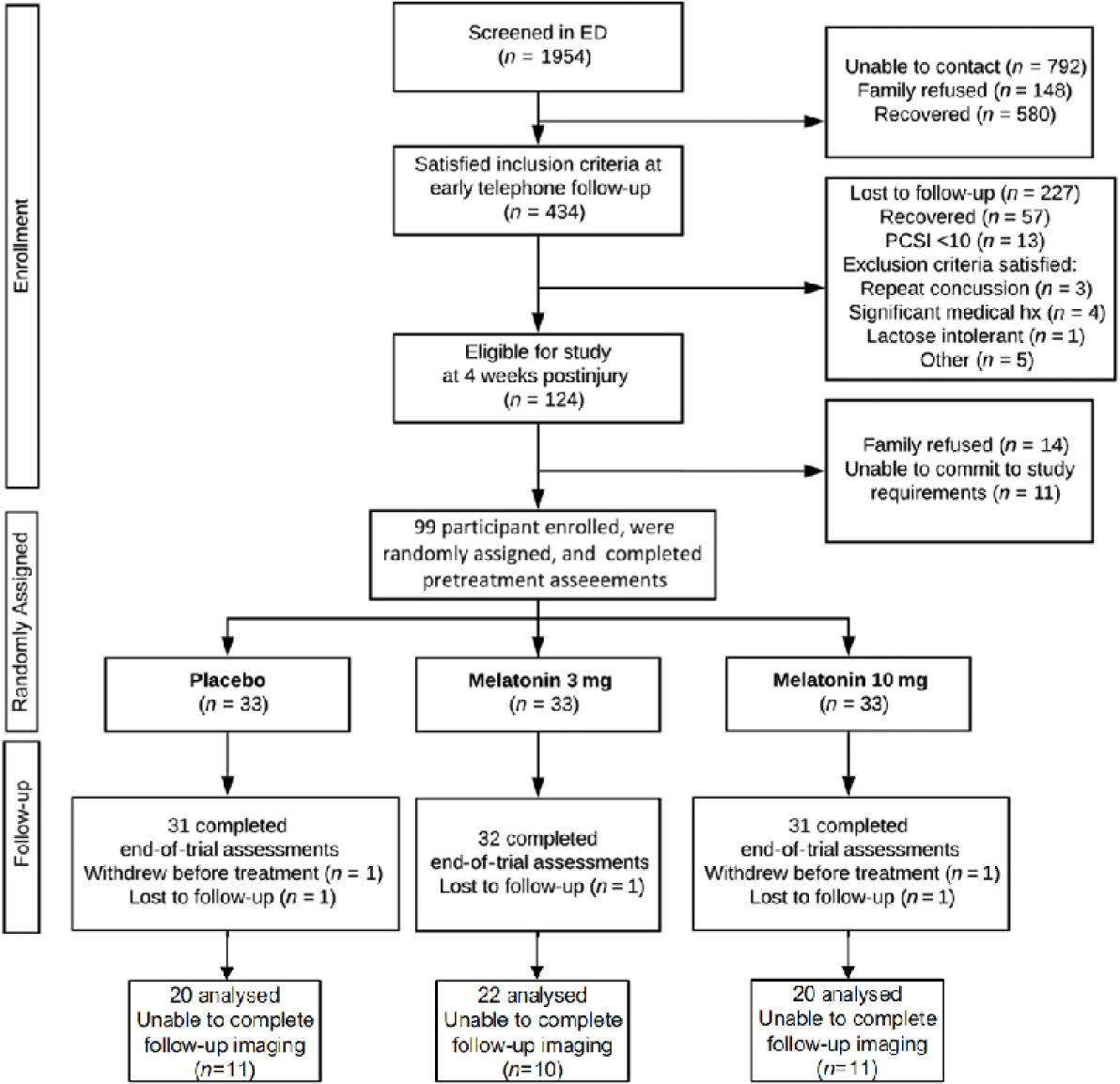
Study consort diagram. The final sample with neuroimaging scans available at both pre and post study points, comprising of 62 participants randomly split across placebo, 3mg melatonin and 10mg melatonin groups

### Outcome measures

The following clinical and behavioral measures were considered for this study:

i. The primary outcome was change in Post-Concussion Symptom Inventory total score for youth self-report (PCSI-Y), calculated as pre-treatment (4 to 6 weeks post-injury) minus end-of-treatment (8 to 10 weeks post-injury) score. The PCSI consists of 26-item questionnaires used to summate key domains of post-concussive symptoms and behaviors.^26^ These domains include physical, cognitive, emotional and sleep/fatigue. As reported elsewhere,^25^ participants were considered to have *recovered* if the post-treatment total PCSI score returned to preinjury levels or below (assessed at enrollment using pre-injury PCSI) and they had returned to normal activities.
ii. Sleep-wake disturbance scores were calculated using the sleep/fatigue domain questions of the PCSI assessing trouble falling asleep, sleeping more than usual, sleeping less than usual, drowsiness and fatigue. Each symptom was rated using a Guttmann scale, 0 (not a problem) to 6 (almost always a problem). These five sleep-related questions were summed to obtain a PSCI “total sleep score” (maximum possible score of 30). It is noted that change in total PCSI (minus sleep) and change in sleep scores were related, i.e. decreases in PCSI correlated with sleep improvement (*r* = −0.57, *p* <0.001).
iii. Actigraphy data was also collected to obtain objective sleep activity behavior over the treatment period using the Actiwatch-2 (Philips Respironics). Pre-treatment sleep parameters from this cohort have been recently reported.^27^ To summarize, the Actiwatch-2 was worn continuously on the non-dominant wrist for 5 to 7 days, using 15 second data epochs. Data analysis was performed using Respironics’s Actiware analysis software (version 6.0.2), where “sleep” or “wake” periods were scored according to weighted movement activity above and below wake thresholds.^27^ Children in the study kept a self-reported sleep diary which documented bedtime, sleep time, wake time, number and length of naps, medications used, watch removal, and number of caffeine-containing beverages taken (if any). Actigraphy data was validated against information provided in these sleep diaries, and potential artifacts were excluded (e.g. watch removal, as confirmed via a subject’s sleep diary or when periods of zero activity counts for greater than ten minutes were recorded). Actigraphy measures included: (i) change in Wake After Sleep Onset (WASO): this measure constituted the total number of estimated minutes between sleep onset and the end of a sleep period that was scored as “wake” by the sleep/wake analysis; and (ii) total sleep time: this was the calculated period of time in minutes from sleep onset and sleep end. These measures were used as objective measures of sleep behavior before and after treatment periods. Of note, changes in subjective sleep scores from PCSI were not significantly correlated with changes in WASO (*r =* 0.07, *p =* 0.64) nor total sleep time (*r =* 0.06, *p =* 0.30). Changes in WASO were correlated with total sleep time (*r* = 0.30, *p* = 0.046).

### Neuroimaging

Neuroimaging was obtained the day before treatment commenced and repeated during the week following treatment. A 3.0 T GE scanner (Healthcare Discovery MR, 750w, 32-channel head coil) was used to acquire structural (T1) and resting-state functional (rs-fMRI) images. Structural scans (T1-weighted) were acquired in the oblique axial plane via the following parameters: 0.8 mm slice thickness, flip angle = 10°, inversion time = 600 ms, FOV = 240 mm. Echo-planar images (rs-fMRI) were collected for five minutes and ten seconds (2000 ms TR, 30 ms TE, 90° flip angle, 3.6 mm slice thickness, 23 × 23 cm FOV, 64 × 64 matrix). As reported elsewhere,^9, 21^ pre-processing included regression of nuisance covariates including head motion (Friston 24), linear trends, and signals from cerebrospinal fluid (CSF) and white matter (WM). Our final cohort comprised of a total of 62 subjects, after excluding 6 subjects due to absence of post-treatment scans or failure to pass quality control measures. See **Table 1** which summarizes demographic and neuroimaging measures for each of the treatment groups. Neuroimaging analyses were performed within MATLAB (MathWorks, Natick, Massachusetts), using SPM12 and custom scripts. Visualizations for our figures were generated using a combination of MATLAB, MRIcroGL and online tools (**Fig. 2**: https://github.com/ihstevenson/beeswarm; Beeswarm plot; **Fig. 3**: http://immersive.erc.monash.edu.au/neuromarvl/; Connectogram).

**Table 1.** Subject demographics and neuroimaging summary characteristics across treatment groups. Sample size, age and gender distribution for the three treatment groups averaged across pre- and post-treatment periods are summarized, with median and standard deviation (SD) where appropriate. Quality control neuroimaging measures including total intracranial volume and grey matter (GM); along with framewise displacement measurements (FD) from rs-fMRI are reported. Changes in sleep measures from sleep scores from the PCSI subscale and wrist actigraphy (where negative values indicate improvement), pre to post treatment are also provided. No significant differences were observed for these sleep measures across melatonin treatment groups. No significant differences were observed across treatment groups for demographic and neuroimaging measures: Chi Square test assessed group differences in Age and Gender whereas Wilcoxon rank sum test assessed for differences in quality control measures.

| Sample characteristics | Group summary |  |  |
| --- | --- | --- | --- |
| **Demographic** | *Placebo*<br>(n=20) | *Melatonin*<br>3 mg (n=22) | *Melatonin*<br>10 mg (n=20) |
| Age | 15.2 (1.8) | 13.5 (2.8) | 14.3 (2.7) |
| Gender *Male / female* | 7/13 | 10/12 | 10/10 |
| **Change in sleep measures** |  |  |  |
| Sleep score (PCSI-subscale) | -3.60 (4.45) | -2.64 (3.22) | -2.35 (4.61) |
| WASO (minutes) | 7.43 (11.51) | -4.58 (11.58) | -2.88 (9.15) |
| Total Sleep time (minutes) | -15.0 (84.78) | -26.0 (48.80) | -14.47 (41.60) |
| **Anatomical measures** |  |  |  |
| Estimated total intracranial volume (TICV, cm<sup>3</sup>) | 1485.0 (149.4) | 1407.5 (111.3) | 1482.5 (118.5) |
| Grey Matter (GM, cm<sup>3</sup>) | 783.6 (80) | 794.3 (62.4) | 790.0 (76.2) |
| **Functional measures** |  |  |  |
| Frame-wise displacement, FD Power (mm) | 0.15 (0.06) | 0.14 (0.06) | 0.14 (0.05) |

**Figure 2.**
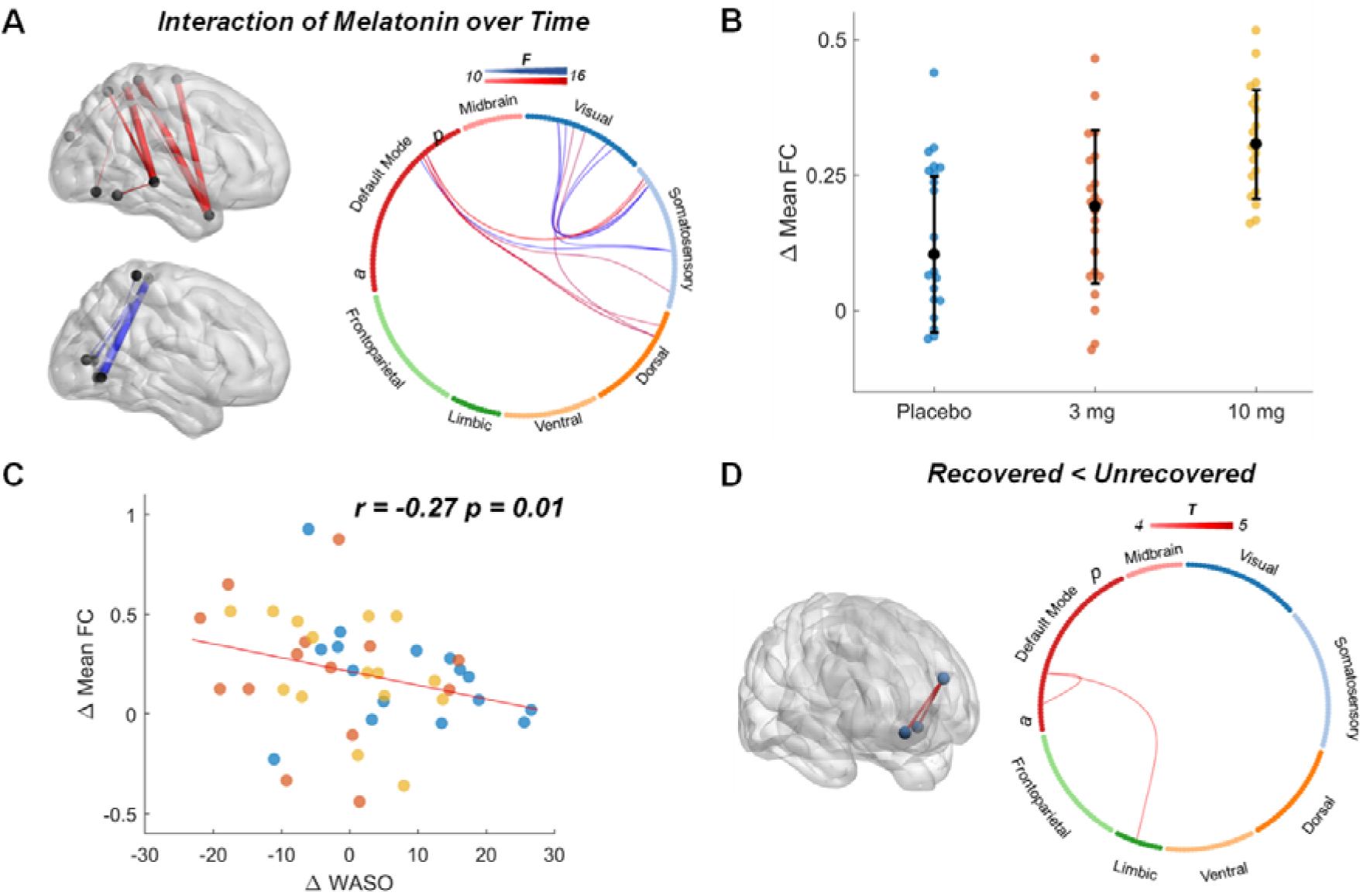
Changes to whole-brain FC following melatonin treatment in pediatric PPCS. **A)** Brain networks showing a melatonin over time interaction. Labels ‘*a*’ and ‘*p*’ indicate anterior and posterior regions of the DMN. The two sub-networks, as detected by NBS, are highlighted in red (sub-network 1) and blue (sub-network 2). **B)** Change in mean FC (post minus pre) of all network edges across all subjects in each treatment group, where black error bars indicate the mean ± standard deviation (SD). Individual subjects are allocated by treatment groups indicated by specific colors where placebo = blue, 3 mg = orange and 10 mg = yellow. **C)** Change in mean FC values negatively correlated with WASO measures, where higher FC post-treatment corresponded with reduced WASO. **D)** Network contrasts of PPCS patients who did not recover (*n* = 39) had higher FC in anterior DMN and limbic nodes versus patients who did recover (*n =* 23) following the treatment period (*p*_FWE_ < 0.05, *T*_*2,61*_ = 4.3). All networks adjusted for age and gender.

**Figure 3.**
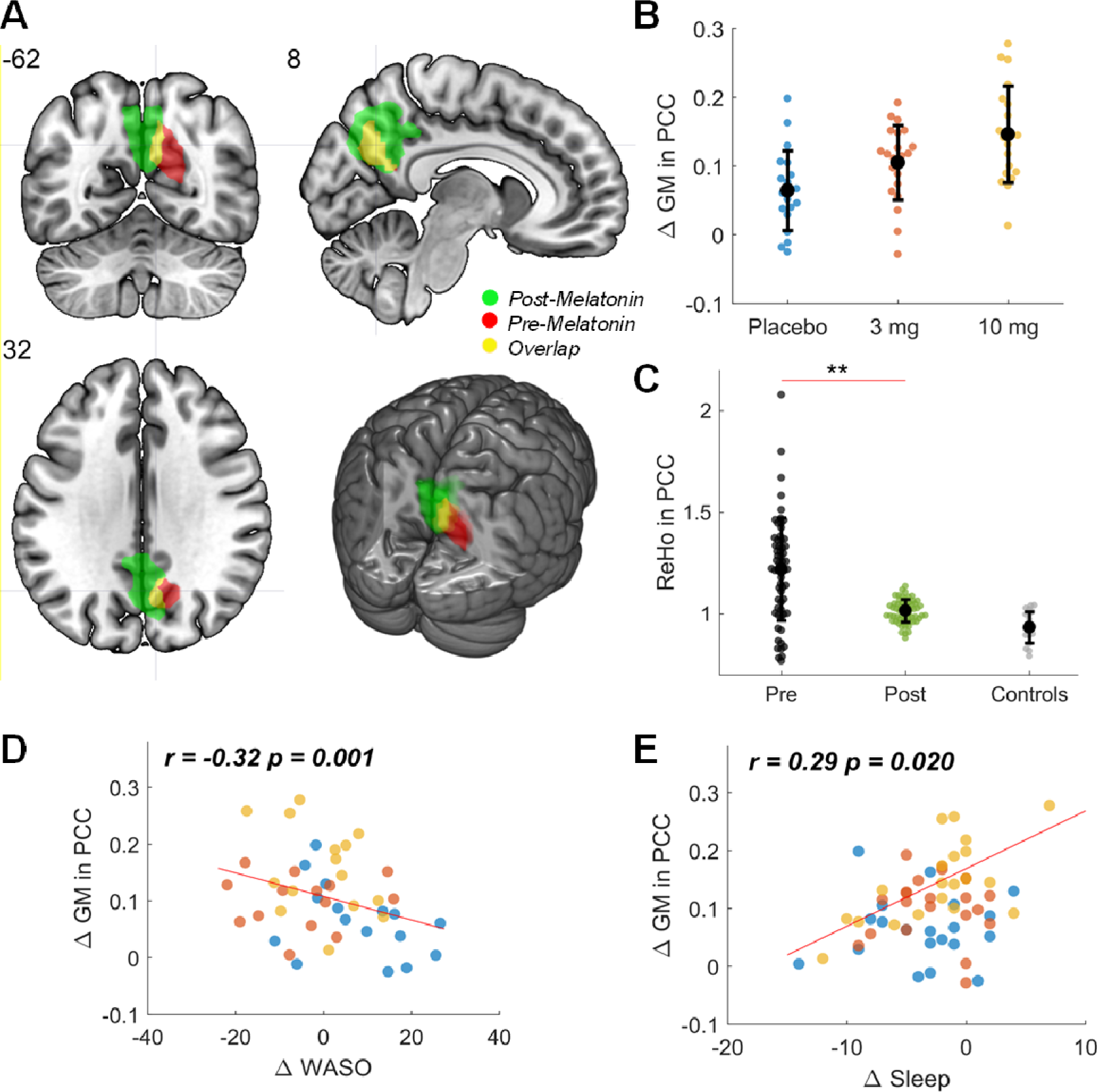
Changes to whole-brain grey matter (GM) and intra-regional connectivity (ReHo) within the PCC following melatonin treatment in children with PPCS. **A)** Pre-melatonin GM clusters in the PCC (indicated in red, from our previous study^9^) were identified from whole-brain analyses to be negatively correlated with sleep and fatigue disturbances. The main effect of group of whole-brain GM post-melatonin in this study is also shown (indicated in bright green). The overlap (indicated in yellow) highlights a mutual region of change. **B)** Changes in PCC GM contrasted across melatonin treatment groups, where black error bars indicate mean ± SD. **C)** ReHo estimates across pre and post treatment periods, with black and green dots respectively. A healthy control cohort (*n* = 20, indicated in grey) from our previous study^9^ have been included as a comparator. Double asterisks indicate *p*_*uncorr*_ < 0.001. **D)** PCC GM changes negatively correlated with change in WASO, with higher GM corresponding with reduced WASO. **E)** Higher PCC GM change positively correlated with increased sleep post-treatment. Individual subjects are allocated by treatment groups indicated by specific colors where placebo = blue, 3 mg = orange and 10 mg = yellow. All contrasts adjusted for age and gender.

Following earlier work,^9,21^ we measured functional connectivity (FC) using two methods. We first examined whole-brain FC using a validated parcellation comprising 214 regions.^28^ Whole-brain FC was also calculated by computing Pearson correlations in rs-fMRI signals across all pairs of brain regions and connectivity matrices were derived. A Fisher’s r-to-z transformation was applied to all connectivity matrices to normalize values. A Network Based Statistic (NBS) approach^29^ was used to identify changes in whole-brain FC across treatment groups. NBS offers an equivalent approach to cluster-level thresholding of statistical parametric maps, by accounting for the multiple comparisons problem that occurs for numerous univariate tests of connectivity. Further, for a given statistical contrast (e.g. paired t-test) it determines the extent to which interconnected set of connections are related to the effect of interest. In doing so, differences in network topologies (sub-networks) related to specific groups can be derived. In addition to whole-brain FC, the second method we used to examine intra-regional FC was via analyzing regional homogeneity (ReHo). ReHo is a voxel-based measure of temporal synchronization of the BOLD signal between a small set of spatially adjacent neighboring voxels. Whole-brain ReHo estimates were derived from unsmoothed rs-fMRI data by calculating the Kendall coefficient of concordance and voxel cluster size of 27.

We also assessed grey matter volumes by performing voxel-based morphometry (VBM) on T1 scans. VBM is a fully automated whole-brain measurement of region-specific grey matter volumes within a subject’s T1 scan. We operationalized our VBM processing pipeline via the SPM12 toolbox. Full details of our VBM pipeline are specified in our prior study.^9^ To summarize VBM processing steps, grey matter volumes are estimated via (i) spatial normalization and age-based template-matching of tissue probabilities, brain size and cortical thickness across child and adolescent populations^30, 31^ (ii) segmentation of brain volumes to extract grey matter (GM), CSF and WM images (iii) diffeomorphic anatomical registration^32^ to optimize volume correction and inter-subject alignment across images; and finally (iv) smoothing and modulation of images via normalization, where images were smoothed using an isotropic Gaussian kernel at 9 x 9 x 9 mm full-width at half maximum (FWHM) and voxel sizes of 3 x 3 x 3 mm for spatial normalization. For quality control, statistical contrasts of total intracranial brain volume (TICV) and GM across treatment groups were performed to ensure that differences were not driven by these anatomical measures (reported in **Table 1**).

### Statistical analyses

For whole-brain FC, ReHo and GM measures, we adopted a longitudinal ANCOVA model. This model tested each neuroimaging measure independently to assess the interaction between treatment groups (placebo, 3mg melatonin, and 10 mg melatonin) and time (pre-treatment, post-treatment) covarying for age and gender.

Statistical contrasts were setup within NBS and SPM toolboxes. ANCOVA outputs were used to inform further statistical analyses alongside our sleep measures. For interaction effects within the ANCOVA model, our null hypothesis was that FC in melatonin treatment groups would not differ to placebo over time (*H*_0_: ΔFC_10mg *melatonin*_ = ΔFC_3mg *melatonin*_ *=*ΔFC _placebo_), whereas our alternative hypothesis put forward that functional connectivity would positively change with melatonin over time (*H*_α_: ΔFC _10mg *melatonin*_ > ΔFC _3mg *melatonin*_ > FC _*placebo*_). For determining statistical significance in whole-brain FC, 5000 permutations were applied and only components that survived a network-level threshold of *p*_FWE_ < 0.05 were deemed significant. For ReHo and GM measures, FWE correction was performed at cluster-level (*p*_FWE_ < 0.05, search threshold of *p*uncorr < 0.001).

Following the results of our ANCOVA model, we then independently tested for correlations between change in whole-brain FC, ReHO, and GM measures and change in sleep and WASO measures. Change was calculated as end-of-treatment minus pre-treatment. For our measures, we performed partial correlations from pre-treatment imaging with WASO to account for any potential confounders to our observations.

Differences between pre and post-treatment groups in those children who recovered from PPCS versus those children that did not fully recover (i.e. remained symptomatic) were also examined. An unpaired t-test was employed to examine post-treatment differences between recovered versus unrecovered groups. For this contrast, our null hypothesis was that, independent of study drug, those subjects who recovered from PPCS would not differ to those who did not recover (*H*_0_: FC _recovered_ = FC _unrecovered_), whereas our alternative hypotheses tested for decreases (*H*_α1_: FC _recovered_ > FC _unrecovered_) and increases (*H*_α2_: FC _recovered_ < FC _unrecovered_) in our measures for those children who remained symptomatic.

## RESULTS

Results showed a significant group by time interaction in whole-brain FC in two sub-networks (**Fig. 2A**, *p*_FWE_ *=* 0.03, *F*_*2,61*_ > 10, Cohen’s *d* = 0.87). Sub-network 1 consisted of edges between posterior DMN with visual, somatosensory, and dorsal areas (indicated by red edges, **Fig. 2A**) while sub-network 2 consisted of edges within visual and somatosensory regions (highlighted by blue edges, **Fig. 2A**). The change in mean FC (post minus pre-treatment) of sub-network edges (all edges collapsed), across all participants indicated increasing FC for those treated with melatonin (**Fig. 2B**). In assessing correlations between global (all groups collapsed) change in FC and sleep measures, we found that change in mean FC negatively correlated with change in WASO (**Fig. 2C**, *r =* −0.27, *p*_*uncorr*_*=* 0.01) but was not associated with total sleep time (*r =* 0.07, *p*_*uncorr*_ = 0.43). Adjusted *r*-values for the WASO relationship indicates the relative contribution of FC in participants treated with melatonin compared to placebo (*r*_*melatonin*_ *=* −0.30, *r*_placebo_ *=* −0.13). Importantly, the correlation between change in FC and WASO across all treatment groups was not driven by pre-treatment FC (partial correlations between pre-treatment FC and WASO revealed no significant associations, *r =* 0.03, *p*_*uncorr*_ = 0.84). Change in mean FC across all treatment groups was not significantly correlated with overall improvement in sleep-related problems (*r =* 0.17, *p*_*uncorr*_ = 0.18).

In comparing recovery groups, increases in post-treatment FC between the anterior DMN nodes (medial prefrontal cortex) and a limbic node (paralimbic orbitofrontal area, **Fig. 2D**, *p*_FWE_ *=* 0.026) were detected in participants who did not recover from PPCS. No significant decreases in FC and ReHo between recovery groups were detected.

In assessing changes in whole-brain GM volume between melatonin groups versus placebo, a main effect of group was detected, where significant increases in GM within the PCC (**Fig. 3A**) for post-treatment subjects were found compared to subjects prior to treatment (*p*_FWE_ = 0.0013, cluster-level (*k*_E_) ≥ 533 voxels, *F*_*2,61*_ = 21.12, Cohen’s *d* = 1.27). No group by time interactions were detected. However, Plotting the change in mean PCC GM (post minus pre-treatment) across all groups highlighted increasing GM for participants treated with melatonin, as well as a dose effect (**Fig. 3B**).

No significant effects were found when examining the interaction of melatonin over time for whole-brain ReHo findings. No significant main effect of group was detected for ReHo across subjects (*p* = 0.10, *F*_*2,61*_ = 2.4). Correlations between ReHo and all our sleep measures (sleep scores and actigraphy) were also non-significant. However, a main effect of time was found for estimates of ReHo in the PCC, where post-treatment groups were significantly different to pre-treatment subjects (one-way ANOVA, *p* = 1.7 x 10^−7^, *F*_*2,61*_ = 36.30, **Fig. 3C**).

Changes in PCC grey matter were also correlated with sleep measures, where GM increases in this brain region corresponded with reduced WASO (*r* = −0.32, *p*_*uncorr*_ *=* 0.001, **Fig. 3D**) and improved sleep-related problems (*r* = 0.29, *p*_*uncorr*_ *=* 0.02, **Fig. 3E**). Adjusted *r*-values for these correlations indicated the relative contribution of GM in participants treated with melatonin compared to placebo (for WASO: *r*_*melatonin*_ *=* −0.28, *r*_placebo_ *=* −0.17 and; improved sleep-related problems: *r*_*melatonin*_ *=* 0.24, *r*_placebo_ *=* 0.18). Results showed that partial correlations of pre-treatment GM with WASO revealed no significant associations (*r =* 0.04, *p*_*uncorr*_ = 0.14). No significant correlation was detected for GM changes with total sleep time (*r* = 0.10, *p*_*uncorr*_ = 0.23). Additionally, no significant differences in whole-brain GM was found when comparing children with PPCS who recovered post-treatment (*n =* 23) versus those that did not recover (*n =* 39).

## DISCUSSION

Our results showed that a 4-week course of melatonin treatment in children with PPCS contributed to dosage-related modulations of functional connectivity and structure in defined brain networks. Melatonin-related changes in structure and function were correlated with improvement in subjective and objective sleep parameters. Specifically, children with PPCS that reported better sleep and reduced wake periods following sleep onset demonstrated a net increase in grey matter within the PCC. These grey matter changes corresponded with overall increases in functional connectivity between posterior DMN nodes and visual, somatosensory and dorsal brain networks. Those children that did not recover from PPCS post-treatment had significantly increased functional connectivity between anterior DMN and limbic network regions. Taken together, these results suggest that children with PPCS may benefit from melatonin treatment as there are discernable changes to core brain functions interacting within and between DMN regions. Despite the absence of a direct clinical benefit, findings from this study provide some evidence supporting a protective effect of melatonin on brain function and structure in children presenting with persistent post-concussion symptoms. Although speculative, this could be due to brevity of the course of treatment (4 weeks), other non-measured non-injury related factors that influence symptom reporting, and/or insensitivity of the clinical outcome measures.

This is the first pediatric mTBI study to provide evidence linking changes in brain function and structure before and after a pharmacological treatment, melatonin. Our results reveal important associations between resting-state networks of DMN, visual and somatosensory regions in children with PPCS following melatonin treatment. Relatively recent neuroimaging studies have shown that compromises to the brain’s structure-function relationships, such as the DMN, have been associated with increased sleep disturbance and fatigue following mTBI,^9, 33, 34^ which may respond to exogenous melatonin. In this regard, melatonin treatment is associated with promoting sleep-like changes in posterior DMN regions including increased activation within the precuneus.^35, 36^ Our study supports these assertions, by showing that the effect of melatonin treatment does increase grey matter specifically within the PCC (**Fig. 3A**), and that these increases are dose-related (**Fig. 3B**). We also observed a change in ReHo values (**Fig. 3C**) within the PCC, though this seems to suggest recovery related effect over time rather than an effect of melatonin treatment. Our findings of grey matter and ReHo change are in line with the broader role of the PCC in health and disease,^37^ and suggests there is a compensatory effect occurring within this region so that normal functions in these children are reassuringly restored post-injury.

Whilst mechanisms of action linked to the administration of melatonin, such as decreased inflammatory response, may explain the positive effect on brain activity, it is likely that dose-related responses in brain structure and function are more closely linked to mechanisms of synaptic plasticity and improved regulation of circadian rhythms.^38-40^ Though the latter has been well studied with respect to melatonin effects, there is convergent evidence from neuroimaging and preclinical data to suggest that increases in sleep periods and reduced overnight wake periods contributes to increases in synaptic strengthening, neuronal firing, and whole-brain network coherence.^39, 41^ Our findings highlight the importance of understanding the neurophysiological factors underpinning the observed effects and suggest that a closer look into brain activity during overnight sleep patterns may provide critical insights into brain network reorganization in concussed children with persistent sleep-related disturbances.

Whilst only a few studies have examined TBI-related sleep disturbances and long-term functional connectivity changes during recovery,^34, 42^ resting-state functional connectivity studies in children and youth have further elucidated why the DMN plays a key role in mediating sleep behaviors.^17, 43-45^ Such studies have shown that younger cohorts with early “chronotypes” (i.e. behavioral traits of sleep, where early chronotypes have early sleep onset and better maintenance of lifestyle activities) have increased functional connectivity between DMN and salience networks, which in turn contribute to better cognitive control and overall sleep function.^44^ Another study found that in young adults with later chronotypes (i.e., those with later sleep onset at night and poorer maintenance of lifestyle activities) had decreases in connectivity within posterior DMN nodes, which contribute to higher vulnerability for depression. Though our study did not classify chronotypes and sleep behaviors specifically, there were observable inter-network increases in functional connectivity between posterior DMN nodes with visual, somatosensory and dorsal regions which correlated with reduced WASO periods post-treatment. Importantly, these findings did not significantly covary with gender or age, which suggests that functional connectivity changes reflect mechanisms of injury, given that adolescent populations are, in general, typically more prone to sleep and chronotype disturbances.^46^ Our results suggest that children recovering from PPCS may be more amenable to changes in functional connectivity between DMN and other key brain regions following melatonin administration, contributing to more widespread network activity and improvement in sleep behaviors.

There are some important limitations that require consideration. Firstly, determining effective treatments that resolve PPCS remains an open question. Although melatonin treatment identified neural correlates that corresponded with improved sleep and recovery in some children with PPCS, those children that remained symptomatic had notably increased functional connectivity within anterior DMN nodes and limbic network regions in the post-treatment period (**Fig. 2D**). Slower recovery rates observed in those children that remain symptomatic could reflect the association of hyperconnectivity within anterior DMN nodes^47^ and a dysregulation of neuronal activity from thalamic regions.^48^ In this regard, further longitudinal studies are required to ascertain whether these connectivity patterns normalize over time in youth with PPCS. Secondly, our study did not employ a validated pediatric sleep questionnaire to assess sleep-related problems, but instead used relevant questions from a well validated concussion symptom questionnaire (PCSI), which evaluated the key areas of sleep concerns for our population (i.e., fatigue, drowsiness, trouble falling or staying asleep), but did not assess pre-sleep arousal symptoms. Though the PCSI has demonstrated moderate to strong internal consistency (0.79) and test-retest reliability (0.69 to 0.73),^49^ the use of well validated specific sleep questionnaires could potentially offer better sensitivity in characterizing sleep patterns and disturbances in these types of pediatric cohorts.^50^ Lastly, though the inclusion and matching of actigraphy measures with sleep diaries in this cohort significantly improves the interpretation of the findings, measures such as WASO may underestimate the overall fragmentation of sleep periods measured (e.g. the child may be in a wake state in bed without moving and thus limiting detection on wrist actigraphy), however this is likely to be independent of treatment group or time post-injury.

In summary, for children experiencing increased sleep disturbances and fatigue following PPCS, the administration of melatonin may assist with compensatory recovery of brain functions namely in regions interacting with, but not limited to, the DMN. Whilst the treatment of melatonin did not result in overall recovery from PPCS, it did have a positive behavioral effect in improving sleep parameters. Thus, our work motivates future studies examining the long-term benefits of melatonin treatment, and improved sleep, to overall PPCS recovery. These studies could consider investigating low-risk treatments with a combination of targeted, personalized interventions such as: (i) non-invasive brain stimulation to selectively modulate functional connectivity between DMN and prefrontal brain regions to reduce PPCS symptoms (e.g. working memory, mood-related problems); and/or (ii) symptom directed therapy (e.g. cognitive behavior therapy for sleep disturbance and insomnia in post-concussion syndrome^44^) for those children whom report higher fatigue, pain, anxiety and cognitive difficulties.

## Data Availability

The dataset will be made available upon request to the principal investigator (K.M.B). The use of this dataset in further scientific work will require a data sharing agreement with the University of Calgary.

## ACKNOWLEDGEMENTS

K.M.B. acknowledges funding support from the Canadian Institutes of Health Research (grant number: 293375) and Motor Accident Insurance Commission (MAIC, Queensland, Australia). L.C. is supported by the Australian National Health Medical Research Council (L.C. 1099082 and 1138711).

